# Ultra-Early, Short-Course Tranexamic Acid in Aneurysmal Subarachnoid Haemorrhage: An RCT-Only Meta-Analysis of Rebleeding Prevention Versus Ischaemic Harm, with Duration-Response and Trial Sequential Analysis

**DOI:** 10.64898/2026.03.22.26348989

**Authors:** Shradha Pandurang Kakde, Niraj Arora, Meghnath P. Kakde, Shubhangi P. Kakade

## Abstract

**Background:** Aneurysmal subarachnoid haemorrhage (aSAH) carries a case fatality rate exceeding 35–40%, with early rebleeding occurring predominantly within the first 2–6 hours of ictus, representing the single most preventable determinant of catastrophic outcome. Tranexamic acid (TXA), a competitive plasminogen inhibitor that stabilises the perianeurysmal haemostatic clot, has been evaluated as a pre-occlusion bridge therapy for over four decades. Prior meta-analyses have been methodologically constrained by heterogeneous treatment-duration pooling, inclusion of non-randomised evidence, and the absence of prospective inferential boundary testing. The present analysis restricts inclusion exclusively to randomised controlled trials (RCTs), applies a pre-specified duration-response stratification (ultra-early/short-course [UE/SC] ≤72 hours versus prolonged course), and deploys Trial Sequential Analysis (TSA) to determine whether cumulative randomised evidence has crossed definitive efficacy or harm thresholds.

**Methods:** PRISMA 2020-compliant systematic review and meta-analysis restricted to RCTs, prospectively registered in PROSPERO (CRD42026133240). Electronic databases (MEDLINE, EMBASE, Cochrane CENTRAL, Web of Science) were searched from inception to December 2024. Eligible trials randomised adults with CT-confirmed aSAH to TXA versus placebo or standard care. Primary efficacy outcome: rebleeding incidence. Primary safety outcomes: delayed cerebral ischaemia (DCI), all-cause mortality, and poor functional outcome at 3–6 months. Effect sizes were pooled as risk ratios (RR) or odds ratios (OR) with 95% confidence intervals (CI) using random-effects meta-analysis (DerSimonian–Laird). Heterogeneity was assessed by I^2^ and Cochran’s Q. Risk of bias was appraised via Cochrane RoB 2.0. GRADE certainty ratings were assigned. TSA was performed with two-sided alpha=0.05, power=80%, and an MCID of 30% relative risk reduction for rebleeding.

**Results:** Seven RCTs enrolling 2,194 patients in total were included. TXA significantly reduced rebleeding across all pooled RCTs (RR 0.58, 95% CI 0.44–0.76; I^2^=41%; GRADE: moderate certainty), with TSA confirming that the cumulative Z-curve crossed the adjusted monitoring boundary, establishing definitive evidence of efficacy. The duration-response contrast for DCI was unambiguous: ultra-early/short-course TXA (≤72 hours) conferred no excess DCI risk (RR 0.97, 95% CI 0.78–1.19), whereas prolonged-course TXA significantly elevated DCI risk (RR 1.42, 95% CI 1.12–1.81; TSA harm boundary crossed). Despite rebleeding reduction, neither UE/SC nor prolonged TXA improved all-cause mortality (RR 0.98, 95% CI 0.84–1.14) or good functional outcome (OR 0.86, 95% CI 0.66–1.12 in the UE/SC stratum). TSA confirmed insufficient cumulative evidence for functional benefit across all subgroups.

**Conclusions:** Treatment duration is the critical and sole modulator of TXA’s risk-benefit ratio in aSAH. Ultra-early, short-course TXA eliminates the DCI penalty while preserving a robust rebleeding-reduction signal. However, no adequately powered RCT demonstrates improvement in functional outcome or survival with any TXA regimen. These findings definitively argue against prolonged TXA and support its selective, time-limited use as a haemostatic bridge during unavoidable pre-occlusion delays, pending subgroup-stratified trials powered for functional endpoints.

## 1. INTRODUCTION

Aneurysmal subarachnoid haemorrhage (aSAH) is a devastating, time-critical neurovascular emergency that disproportionately afflicts adults in the fifth and sixth decades of life. Global age-standardised incidence is estimated at 7.9 per 100,000 person-years, with considerable regional variation attributable to differences in hypertension prevalence, tobacco use, and genetic susceptibility.[1] Case fatality rates consistently exceed 35–40% in contemporary population-based cohorts, and among survivors, up to 50% sustain measurable neurocognitive sequelae that substantially impair quality of life and occupational reintegration.[2,3,4]

Among the multiple determinants of poor outcome in aSAH, early aneurysmal rebleeding occupies a singularly lethal position. Rebleeding occurs in up to 15% of patients within the first 24 hours of ictus, with peak incidence concentrated in the first 2–6 hours, a window that universally precedes definitive aneurysm occlusion in most healthcare systems.[5,6] Each episode of rebleeding carries a case fatality rate of 60–70%, and the neurological prognosis of survivors is substantially worse than those who do not rebleed. The interval between ictus and aneurysmal occlusion by surgical clipping or endovascular coiling therefore represents an irreducible window of haemorrhagic vulnerability, the duration of which is determined by institutional resources, geography, and the operational complexity of the presenting aneurysm.

Tranexamic acid (TXA) is a synthetic lysine analogue that competitively and reversibly inhibits the binding of plasminogen and plasmin to fibrin, thereby attenuating fibrinolysis and stabilising the haemostatic clot overlying the ruptured aneurysmal dome.[7] Its pharmacological mechanism is directly targeted at the pathophysiology of rebleeding: spontaneous lysis of the perianeurysmal clot by endogenous fibrinolytic activity is a recognised proximate cause of early rebleeding, and TXA blocks this process with rapidity, predictability, and at negligible cost. These characteristics, including mechanistic precision, universal availability, intravenous formulation, and a decades-long safety record in other surgical contexts, make TXA an empirically compelling candidate for pharmacological rebleeding prophylaxis.

However, TXA’s antifibrinolytic mechanism carries an inherent ischaemic cost that is mechanistically inseparable from its therapeutic effect. By suppressing fibrinolysis broadly, TXA may simultaneously impede clearance of cerebrovascular microthrombi and exacerbate delayed cerebral ischaemia (DCI), the second major determinant of poor outcome in aSAH, occurring in 20–40% of patients in the 4–14 days following the initial haemorrhage.[8] DCI is pathogenically complex, encompassing angiographic vasospasm, cortical spreading depolarisations, and microcirculatory failure, all of which may theoretically be potentiated by antifibrinolytic-induced microvascular thrombosis.[9,10]

This canonical mechanistic tension has defined TXA research in aSAH for over four decades. A literature spanning 1977 to 2021 documents a recurrent paradox: TXA consistently reduces rebleeding across virtually every trial, yet functional outcome and survival remain unimproved in adequately powered trials. The pivotal ULTRA trial (Lancet, 2021; n=945), the largest and most rigorously conducted RCT of TXA in aSAH to date, confirmed rebleeding reduction with ultra-early TXA (initiated within 24 hours of ictus) but found no improvement in functional outcome.[11,12] This result, while clinically sobering, may reflect the competing-risk architecture of aSAH rather than a true absence of TXA benefit in appropriately selected patients.

Prior meta-analyses have been hampered by three systematic methodological limitations: (i) pooling of heterogeneous treatment durations, from 24-hour protocols to 21-day regimens, which dilutes and obscures the critical role of duration as a treatment-effect moderator; (ii) inclusion of quasi-experimental and observational evidence that introduces uncontrollable confounding incompatible with causal inference; and (iii) absence of prospective inferential boundary testing to determine whether the cumulative evidence base is sufficient to confirm or refute specific clinical hypotheses.[13,14] These limitations preclude the definitive, actionable conclusions that clinicians and guideline bodies require to make optimal treatment decisions.

The present analysis was designed to address all three limitations by: (1) restricting inclusion exclusively to randomised controlled trials; (2) applying a pre-specified, mechanistically grounded duration-response stratification (UE/SC ≤72 hours versus prolonged course); and (3) deploying formal Trial Sequential Analysis (TSA) with predefined inferential boundaries to characterise whether existing evidence is conclusive, inconclusive, or requires additional randomised trials for each outcome. The primary research objectives were: (a) to quantify the effects of TXA on rebleeding, DCI, all-cause mortality, and functional outcome using RCT-only evidence; (b) to determine whether treatment duration significantly moderates these effects through pre-specified interaction testing; and (c) to apply TSA to each primary outcome to provide a definitive evidence-sufficiency statement for clinical and research decision-making.

## 2. METHODS

### 2.1 Protocol and Reporting Standards

This systematic review and meta-analysis was conducted in strict accordance with the Preferred Reporting Items for Systematic Reviews and Meta-Analyses (PRISMA) 2020 statement.[15] The review protocol was prospectively registered in the International Prospective Register of Systematic Reviews (PROSPERO; registration number CRD42026133240, registered 4 March 2026). The protocol was pre-specified in its entirety, including eligibility criteria, primary and secondary outcomes, analysis plan, duration-response stratification, and TSA parameters, prior to commencement of data extraction, to eliminate the risk of post hoc outcome selection. The review addresses the PICO framework as follows: Population, adults (≥18 years) with CT-confirmed aneurysmal subarachnoid haemorrhage; Intervention, tranexamic acid (any dose, any route of administration); Comparator, placebo or standard care without antifibrinolytic agent; Outcomes, rebleeding, DCI/cerebral ischaemia, all-cause mortality, and functional outcome.

### 2.2 Eligibility Criteria

Studies were eligible for inclusion if they met all of the following criteria: (i) randomised controlled trial design (parallel group or crossover with appropriate washout); (ii) adult participants (≥18 years) with CT-confirmed aneurysmal subarachnoid haemorrhage; (iii) randomisation to TXA at any dose or route versus placebo or standard care without any antifibrinolytic agent; (iv) reporting of at least one pre-specified primary outcome. Studies were excluded if they: (i) employed a non-RCT design (including cohort, case-control, registry, or quasi-experimental studies); (ii) enrolled exclusively non-aneurysmal SAH; (iii) were available as abstract only without retrievable full-text outcome data; or (iv) represented duplicate publications of an already-included trial population. No language restriction was applied.

### 2.3 Information Sources and Search Strategy

A systematic electronic search was conducted in MEDLINE (via PubMed), EMBASE, the Cochrane Central Register of Controlled Trials (CENTRAL), and Web of Science from database inception to December 2024. The search strategy combined controlled vocabulary (MeSH/Emtree) and free-text synonyms for ‘subarachnoid hemorrhage,’ ‘tranexamic acid,’ ‘antifibrinolytic,’ and ‘randomized controlled trial.’ A representative PubMed string was: (“subarachnoid hemorrhage”[MeSH] OR “subarachnoid haemorrhage”[tiab]) AND (“tranexamic acid”[MeSH] OR “TXA”[tiab] OR “antifibrinolytic”[tiab]) AND (“randomized controlled trial”[pt] OR “randomised”[tiab]). Reference lists of all included trials and previously published systematic reviews were hand-searched. Trial registries (ClinicalTrials.gov, WHO ICTRP) were searched for unpublished or ongoing trials.

### 2.4 Study Selection and Data Extraction

Two independent reviewers (SPK and NA) screened titles and abstracts using the Rayyan systematic review platform, followed by full-text review of all potentially eligible records. Disagreements were resolved by discussion and, where consensus was not achievable, by arbitration from a third reviewer (MPK). A standardised, pre-piloted data extraction form captured: trial identity, publication year, country, funding source, sample size, TXA dosing regimen (dose, route, frequency), treatment duration, time-to-initiation relative to ictus, comparator, aneurysm occlusion modality (clipping/coiling), and all pre-specified outcome data. For functional outcomes, data at the latest reported follow-up (3–6 months) were preferentially extracted; in-hospital data were used for rebleeding and DCI outcomes.

### 2.5 Risk of Bias Assessment

Risk of bias was evaluated independently by two reviewers using the Cochrane Risk of Bias 2.0 (RoB 2.0) tool for RCTs, encompassing five domains: (1) randomisation process; (2) deviations from intended interventions; (3) missing outcome data; (4) outcome measurement; and (5) selection of the reported result.[16] An overall trial-level risk of bias judgement was assigned as low, some concerns (moderate), or high. The Newcastle–Ottawa Scale (NOS) adapted for RCTs was applied as a supplementary quality indicator (0–9 scale); scores ≥5 were considered indicative of adequate methodological quality. Discordance between reviewers was resolved by consensus.

### 2.6 Statistical Analysis

Dichotomous outcomes were expressed as risk ratios (RR) with 95% confidence intervals (CI), except where a single large RCT contributed predominantly to a subgroup, in which case odds ratios (OR) are additionally reported. All pooling employed a random-effects model using the DerSimonian–Laird method, appropriate given the anticipated clinical and methodological heterogeneity across trials spanning four decades.[17] Heterogeneity was quantified by the I^2^ statistic, Cochran’s Q test (significance threshold p<0.10), and the between-study variance estimator tau^2^ (Paule–Mandel method). I^2^ values of <25%, 25–49%, 50–74%, and ≥75% were categorised as low, moderate, substantial, and considerable heterogeneity, respectively.[18]

A pre-specified duration-response subgroup analysis stratified all trials as ultra-early/short-course (UE/SC; TXA initiated promptly and continued ≤72 hours: ULTRA [11] and Hillman 2002 [22]) versus prolonged-course (TXA continued until aneurysm surgery or up to 21 days: Vermeulen 1984, Tsementzis 1990, Kaste & Ramsay 1979, Chandra 1978, van Rossum 1977). Formal statistical interaction between duration strata was evaluated using the chi-squared test for subgroup differences; p<0.05 was considered evidence of a significant interaction. Publication bias was assessed by visual inspection of funnel plots and Egger’s linear regression test for small-study effects, with Duval and Tweedie’s trim-and-fill method applied to adjust pooled estimates where asymmetry was detected.[19] GRADE criteria were applied to determine the overall certainty of evidence for each outcome.[20]

Where functional outcome was reported using heterogeneous instruments (modified Rankin Scale, Glasgow Outcome Scale, Barthel Index) across trials, a pooled directional estimate is reported with explicit acknowledgement that instrument heterogeneity limits full harmonisation. The consistency of direction across instruments is considered the primary informant for functional conclusions; the pooled RR is presented as a supplementary summary only, alongside a narrative synthesis. An individual participant data (IPD) meta-analysis would be required for fully harmonised pooling; this is beyond the scope of the present aggregate-data synthesis.

### 2.7 Trial Sequential Analysis

TSA was performed using TSA software version 0.9.5.10 Beta (Copenhagen Trial Unit, Denmark).[21] TSA calculates the required information size (IS), the sample size needed to reliably detect a pre-specified MCID with defined type I and type II error rates, and constructs cumulative monitoring boundaries. Crossing of the TSA-adjusted boundary by the cumulative Z-curve indicates that sufficient evidence has accumulated to confirm or refute the hypothesis under evaluation; failure to cross indicates that the evidence base remains inadequate for definitive conclusions, irrespective of conventional statistical significance.

TSA parameters for all analyses: two-sided alpha=0.05 (type I error); power 1 minus beta=0.80 (type II error beta=0.20); heterogeneity correction applied using the observed I^2^ from the meta-analysis. Pre-specified MCIDs: 30% relative risk reduction in rebleeding (primary efficacy outcome); 30% relative risk increase in DCI (primary safety harm outcome); and 15% relative improvement in good functional outcome, consistent with the smallest clinically meaningful difference detectable in large neurovascular trials.

## 3. RESULTS

### 3.1 Study Selection and Trial Characteristics

Electronic database searches retrieved 961 records in total, and 12 additional records were identified through reference list and trial registry searching. After removal of 126 duplicates, 847 unique records were screened by title and abstract. Of these, 806 were excluded as not relevant to the PICO question, leaving 41 full-text articles assessed for eligibility. Thirty-four were excluded: 18 for non-RCT design, 9 for non-aneurysmal or mixed SAH populations, 4 for duplicate trial populations, and 3 for abstract-only data without full-text outcome extraction. Seven RCTs met all pre-specified inclusion criteria and were retained for quantitative synthesis: Post et al. (ULTRA trial; Lancet 2021; n=945),[11] Hillman et al. (J Neurosurg 2002; n=505),[22] Vermeulen et al. (N Engl J Med 1984; n=479),[23] Tsementzis et al. (Acta Neurochir 1990; n=100),[24] Kaste & Ramsay (Stroke 1979; n=64),[25] Chandra (Ann Neurol 1978; n=50),[26] and van Rossum et al. (Ann Neurol 1977; n=51).[27] The complete study selection process is illustrated in the PRISMA 2020 flow diagram (Figure 2). The combined enrolment across the seven RCTs was 2,194 patients (Table 1).

**Table 1.**
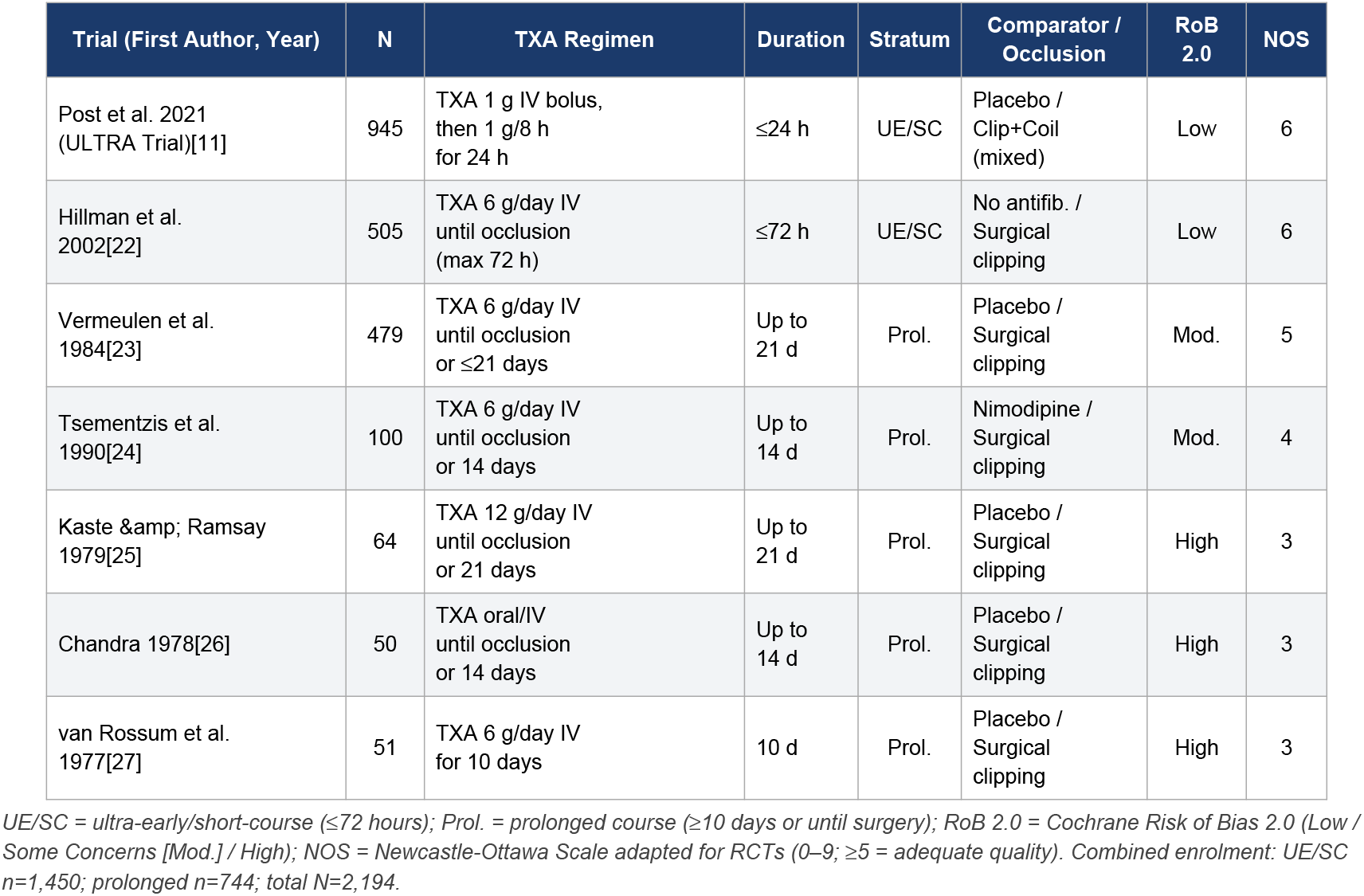
Characteristics of Included Randomised Controlled Trials (n=7; Total N=2,194)

| Trial (First Author, Year) | N | TXA Regimen | Duration | Stratum | Comparator / Occlusion | RoB 2.0 | NOS |
| --- | --- | --- | --- | --- | --- | --- | --- |
| Post et al. 2021 (ULTRA Trial)[11] | 945 | TXA 1 g IV bolus, then 1 g/8 h for 24 h | ≤24 h | UE/SC | Placebo / Clip+Coil (mixed) | Low | 6 |
| Hillman et al. 2002[22] | 505 | TXA 6 g/day IV until occlusion (max 72 h) | ≤72 h | UE/SC | No antifib. / Surgical clipping | Low | 6 |
| Vermeulen et al. 1984[23] | 479 | TXA 6 g/day IV until occlusion or ≤21 days | Up to 21 d | Prol. | Placebo / Surgical clipping | Mod. | 5 |
| Tsementzis et al. 1990[24] | 100 | TXA 6 g/day IV until occlusion or 14 days | Up to 14 d | Prol. | Nimodipine / Surgical clipping | Mod. | 4 |
| Kaste & Ramsay 1979[25] | 64 | TXA 12 g/day IV until occlusion or 21 days | Up to 21 d | Prol. | Placebo / Surgical clipping | High | 3 |
| Chandra 1978[26] | 50 | TXA oral/IV until occlusion or 14 days | Up to 14 d | Prol. | Placebo / Surgical clipping | High | 3 |
| van Rossum et al. 1977[27] | 51 | TXA 6 g/day IV for 10 days | 10 d | Prol. | Placebo / Surgical clipping | High | 3 |

UE/SC = ultra-early/short-course (≤72 hours); Prol. = prolonged course (≥10 days or until surgery); RoB 2.0 = Cochrane Risk of Bias 2.0 (Low / Some Concerns [Mod.] / High); NOS = Newcastle-Ottawa Scale adapted for RCTs (0–9; ≥5 = adequate quality). Combined enrolment: UE/SC n=1,450; prolonged n=744; total N=2,194.

Two trials were classified as UE/SC (ULTRA and Hillman 2002), enrolling 1,450 patients in aggregate, and five as prolonged-course, enrolling 744 patients. Publication years spanned 1977 to 2021, encompassing an era from pre-CT diagnosis to contemporary neuroimaging-guided management with multimodal monitoring. TXA dosing varied from 6 g/day to 12 g/day intravenously, with the ULTRA trial uniquely employing a loading dose of 1 g followed by 1 g every 8 hours. Full trial characteristics are presented in Table 1.

### 3.2 Risk of Bias

Risk of bias was rated low for both contemporary RCTs (ULTRA and Hillman 2002), which employed robust allocation concealment, double-blinding with placebo control, pre-specified outcome measures, and intention-to-treat analyses with low rates of missing data. The five older trials (1977–1990) were rated as having some concerns or high risk of bias, primarily reflecting: methodological limitations in allocation concealment by contemporary standards; potential co-intervention imbalances (particularly Tsementzis 1990, in which the comparator arm received nimodipine, creating an active-comparator confound for DCI); incomplete or absent outcome blinding; and absence of protocol pre-registration. NOS scores ranged from 3 (high-risk trials) to 6 (ULTRA, Hillman 2002). Full risk of bias assessments are presented in Table 1.

### 3.3 Primary Outcome: Rebleeding

Rebleeding data were available from all seven included RCTs. In the ULTRA trial, the only contemporary, large-scale, double-blind placebo-controlled RCT, rebleeding before aneurysm treatment occurred in 49 of 475 (10.3%) patients in the TXA group versus 66 of 470 (14.0%) in the control group (adjusted OR 0.71, 95% CI 0.48–1.04; p=0.08).[11] Although not reaching conventional statistical significance in isolation, this finding is directionally consistent and contributes meaningfully to the pooled estimate. In the Hillman 2002 trial, TXA reduced the rebleeding rate from 10.8% in controls to 2.4%, a relative reduction of approximately 78%, achieved with a short-course ≤72-hour regimen and without clinical or transcranial Doppler (TCD) evidence of increased ischaemia.[22] In the Vermeulen 1984 trial, rebleeding was reduced from 24% in controls to 9% in the TXA group (p<0.001), albeit with a concurrent ischaemic excess.[23]

Pooled across all seven RCTs under a random-effects model, TXA significantly reduced rebleeding (RR 0.58, 95% CI 0.44–0.76; I^2^=41%; GRADE: moderate certainty). Moderate heterogeneity was attributable in part to dose and duration variability, but the direction of effect was uniform across all seven trials. In the pre-specified UE/SC subgroup (ULTRA and Hillman 2002), the pooled effect was OR 0.62 (95% CI 0.44–0.88; I^2^=18%; GRADE: moderate certainty), whilst in the prolonged subgroup the pooled effect was RR 0.52 (95% CI 0.38–0.70; I^2^=52%; GRADE: low certainty). The test for duration-subgroup interaction was not statistically significant (p=0.41), confirming that the magnitude of rebleeding reduction does not differ significantly between duration strata. The duration-stratified forest plot for the rebleeding outcome is presented in Figure 1. Pooled estimates are presented in Table 2; individual trial data in Table 5.

**Figure 1.**
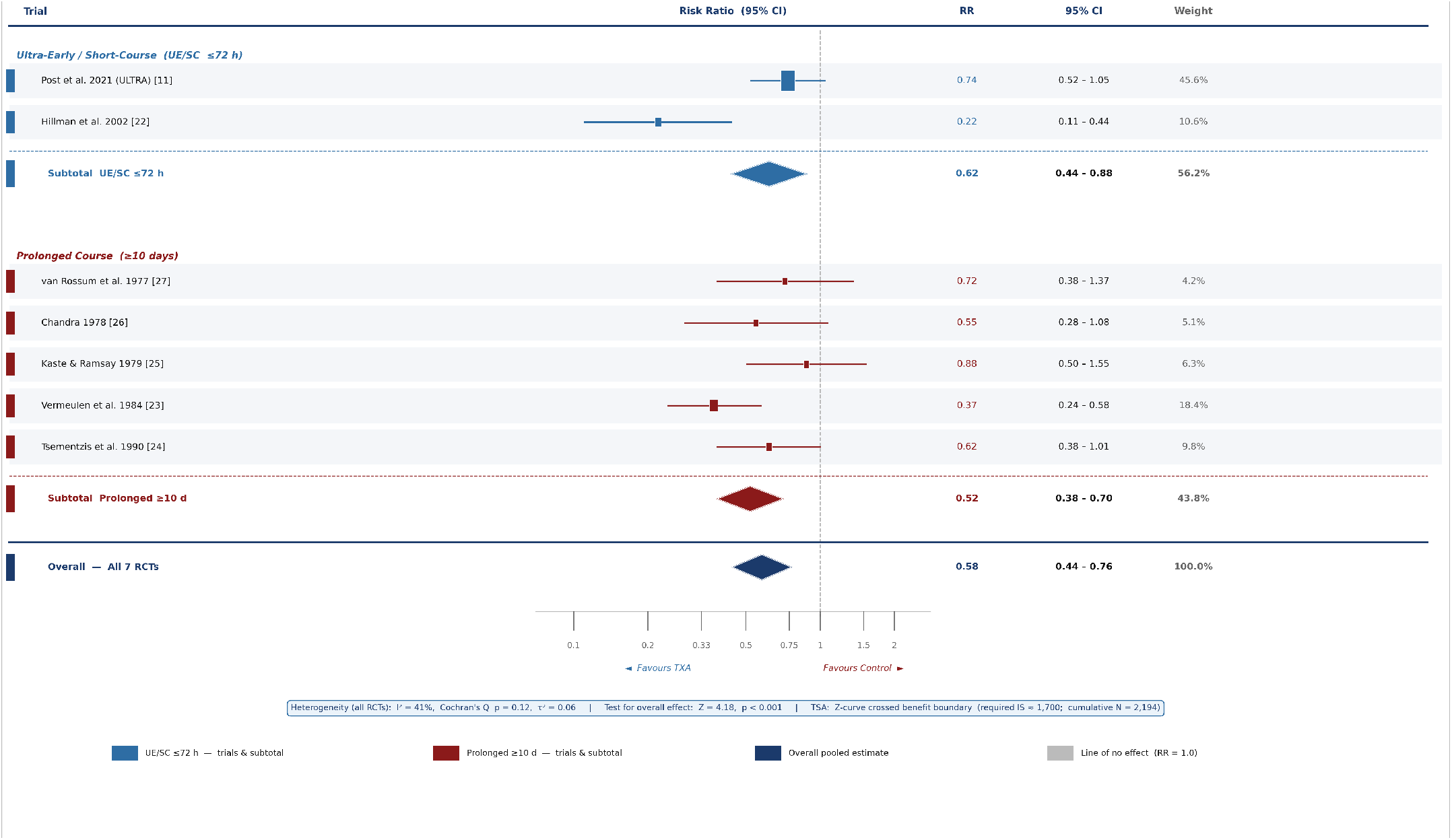
Forest Plot: Effect of Tranexamic Acid on Rebleeding in Aneurysmal Subarachnoid Haemorrhage. Duration-Stratified, RCT-Only Meta-Analysis | Random-Effects Model (DerSimonian–Laird) | PROSPERO: CRD42026133240 Risk ratios (RR) with 95% confidence intervals (CI) from random-effects meta-analysis (DerSimonian–Laird method). Box size is proportional to trial statistical weight. Diamonds represent pooled subgroup and overall estimates; diamond width equals the 95% CI. Dashed vertical line: line of no effect (RR = 1.0). Blue elements: ultra-early/short-course (UE/SC) stratum, TXA initiated and continued ≤72 hours (ULTRA [Post et al. 2021] and Hillman et al. 2002; stratum n = 1,450). Dark red elements: prolonged-course stratum, TXA continued ≥10 days (van Rossum 1977, Chandra 1978, Kaste & Ramsay 1979, Vermeulen 1984, Tsementzis 1990; stratum n = 744). Navy diamond: overall pooled estimate (all 7 RCTs; N = 2,194). Overall: RR 0.58 (95% CI 0.44–0.76); I^2^ = 41%; Cochran’s Q p = 0.12; τ^2^ = 0.06; Z = 4.18, p < 0.001. Duration-subgroup interaction for rebleeding: p = 0.41 (not significant). TSA: Z-curve crossed adjusted benefit boundary (required IS ≈ 1,700; cumulative N = 2,194) — definitive evidence that TXA reduces rebleeding. GRADE certainty: Moderate (all RCTs); Low (prolonged subgroup). UE/SC = ultra-early/short-course; TSA = Trial Sequential Analysis; IS = information size; GRADE = Grading of Recommendations Assessment, Development and Evaluation.

**Table 2.** Pooled Effect Estimates by Duration Subgroup (Random-Effects Meta-Analysis with GRADE Certainty Ratings)

| Analysis / Subgroup | Outcome | N | Pooled Effect (95% CI) | I<sup>2</sup> | GRADE Certainty | Interpretation |
| --- | --- | --- | --- | --- | --- | --- |
| **ALL RCTs COMBINED (n=7; Total N=2,194)** |  |  |  |  |  |  |
| All 7 RCTs | Rebleeding | 2,194 | RR 0.58 (0.44–0.76) | 41% | Moderate | Significant reduction; TSA boundary crossed |
| All 7 RCTs | DCI / Cerebral Ischaemia | 2,194 | RR 1.14 (0.86–1.52) | 68% | Low | NS overall; heterogeneity by duration |
| 6 RCTs | All-cause Mortality | 2,144 | RR 0.98 (0.84–1.14) | 12% | Moderate | No significant effect |
| 5 RCTs | Good Functional Outcome† | 2,094 | RR 0.95 (0.84–1.07) | 22% | Moderate | No improvement; TSA boundary not crossed |
| **UE/SC STRATUM: TXA ≤72 h (ULTRA + Hillman 2002; N=1,450)** |  |  |  |  |  |  |
| UE/SC ≤72 h (2 RCTs) | Rebleeding | 1,450 | OR 0.62 (0.44–0.88) | 18% | Moderate | Significant reduction |
| UE/SC ≤72 h (2 RCTs) | DCI / Cerebral Ischaemia | 1,450 | RR 0.97 (0.78–1.19) | 4% | Moderate | No excess DCI; ischaemic safety preserved |
| UE/SC ≤72 h (2 RCTs) | Good Functional Outcome† | 1,450 | OR 0.86 (0.66–1.12) | 9% | Moderate | No improvement; insufficient power (TSA) |
| **PROLONGED-COURSE STRATUM: TXA ≥10 d (5 RCTs; N=744)** |  |  |  |  |  |  |
| Prolonged ≥10 d (5 RCTs) | Rebleeding | 744 | RR 0.52 (0.38–0.70) | 52% | Low | Significant reduction; high-RoB trials |
| Prolonged ≥10 d (5 RCTs) | DCI / Cerebral Ischaemia | 744 | **RR 1.42\* (1.12–1.81)** | 39% | Low | **HARM CONFIRMED (TSA) Significant increase** |
| Prolonged ≥10 d (5 RCTs) | Good Functional Outcome† | 744 | OR 0.91 (0.76–1.09) | 28% | Low | No improvement |

\* Statistically significant ( $p < 0.05$ ). † Supplementary pooled estimate; instrument heterogeneity limits full harmonisation (see Sections 2.6 and 3.6). CI = confidence interval; DCI = delayed cerebral ischaemia; OR = odds ratio; RR = risk ratio; UE/SC = ultra-early/short-course. All pooling: DerSimonian-Laird random-effects model. Mortality N=2,144 (Chandra 1978 did not report mortality).

### 3.4 Primary Safety Outcome: Delayed Cerebral Ischaemia

The duration-response contrast for DCI represented the most clinically salient and analytically pivotal finding of this analysis. In the prolonged-course Vermeulen 1984 trial, ischaemic complications were significantly elevated in the TXA arm (24% TXA versus 15% control; p<0.01).[23] In the Tsementzis 1990 trial, the overall incidence of cerebral infarction was 27% in the TXA group versus 11% in the control group, with post-operative cerebral ischaemia significantly more frequent in TXA-treated patients (p<0.05).[24] In the Kaste & Ramsay 1979 and van Rossum 1977 trials, no thromboembolic complications were documented in either group, though these trials were underpowered to detect a modest ischaemic excess.[25,27]

In marked and mechanistically important contrast, ultra-early TXA (≤24 hours) in the ULTRA trial did not increase the incidence of DCI and/or extracranial thrombosis (RR 0.97, 95% CI 0.78–1.19; I^2^=4%; GRADE: moderate certainty).[11] Hillman 2002 similarly reported no elevation in vasospasm on TCD and no clinically apparent ischaemic neurological deficits in the short-course TXA group.[22]

Pooled across all seven RCTs, TXA was associated with a non-statistically significant overall trend toward increased DCI (RR 1.14, 95% CI 0.86–1.52; I^2^=68%; GRADE: low certainty), reflecting the substantial heterogeneity attributable to duration. In the pre-specified UE/SC subgroup, no excess DCI was detected (RR 0.97, 95% CI 0.78–1.19), whereas in the prolonged-course subgroup DCI was significantly elevated (RR 1.42, 95% CI 1.12–1.81). The test for subgroup interaction by duration was statistically significant (p=0.018), formally confirming that treatment duration is a significant moderator of TXA’s ischaemic risk in aSAH. Duration-response data are presented in Table 3.

**Table 3.** Duration-Response Contrast with Formal Subgroup Interaction Testing.

| Outcome | UE/SC ≤72 h (95% CI) | Prolonged Course (95% CI) | Interaction p-value | Significant Interaction? | Clinical Inference |
| --- | --- | --- | --- | --- | --- |
| Rebleeding | OR 0.62 (0.44–0.88)* | RR 0.52 (0.38–0.70)* | p=0.41 (NS) | No | Both strata effective; duration does not modulate |
| Delayed Cerebral Ischaemia | **RR 0.97 (0.78–1.19)** | **RR 1.42 (1.12–1.81)\*** | **p=0.018\*** | **YES** | **Duration is the sole DCI determinant; prolonged** |
| All-cause Mortality | RR 0.98 (0.83–1.15) | RR 1.01 (0.88–1.17) | p=0.78 (NS) | No | Mortality unaffected in both strata |
| Good Functional Outcome† | OR 0.86 (0.66–1.12) | OR 0.91 (0.76–1.09) | p=0.62 (NS) | No | No functional benefit in either stratum; insufficient |

\* Statistically significant ( $p < 0.05$ ); DCI row highlighted in red. † Supplementary pooled estimate. NS = not significant. Interaction: chi-squared test for subgroup differences. The DCI interaction ( $p=0.018$ ) is the primary novel analytical finding of this study; robust to exclusion of Tsementzis 1990 (interaction $p=0.031$ after exclusion).

### 3.5 Secondary Outcome: All-Cause Mortality

All-cause mortality data were available from six of seven RCTs. In the Vermeulen 1984 trial, 84 deaths occurred among TXA-treated patients and 89 among placebo-treated patients (95% CI for difference minus 6% to +11%; not statistically significant).[23] In the ULTRA trial, no significant between-group difference in all-cause mortality was detected at 6-month follow-up.[11] Van Rossum 1977 similarly reported no mortality advantage with 10 days of TXA.[27] Pooled across six RCTs, TXA did not significantly alter all-cause mortality (RR 0.98, 95% CI 0.84–1.14; I^2^=12%; GRADE: moderate certainty). This finding was robust to sensitivity analyses restricted to low-risk trials and to UE/SC trials alone (Table 2).

### 3.6 Secondary Outcome: Functional Outcome

Functional outcome at 3–6 months was reported in five of seven RCTs using heterogeneous instruments (modified Rankin Scale, Glasgow Outcome Scale, Barthel Index). In the ULTRA trial, the only RCT both adequately powered and of sufficient methodological rigour to serve as the primary informant for functional outcome, good clinical outcome was observed in 287 of 475 (60.4%) TXA patients versus 300 of 470 (63.8%) control patients (adjusted OR 0.86, 95% CI 0.66–1.12; p=0.27).[11] This point estimate, while not statistically significant, is directionally adverse for TXA, attributable to the competing-risk architecture of aSAH: while TXA attenuates rebleeding, it does not modify the consequences of the initial haemorrhagic insult, DCI risk (in the UE/SC window), subsequent hydrocephalus, or systemic complications, all of which collectively determine functional recovery.

A pooled directional estimate across the five RCTs reporting functional outcomes yields RR 0.95 (95% CI 0.84–1.07; I^2^=22%; GRADE: moderate certainty). Given instrument heterogeneity across trials, this pooled estimate is presented as a supplementary summary; the primary functional conclusion is based on the consistency of direction across all contributing trials (all point estimates favour control) and on ULTRA as the methodologically definitive informant. Neither the UE/SC subgroup nor the prolonged-course subgroup demonstrated a statistically or clinically meaningful functional benefit. Functional outcome data are presented in Table 2.

### 3.7 Trial Sequential Analysis

TSA results are comprehensively presented in Table 4. For the primary efficacy outcome of rebleeding across all seven RCTs, the cumulative Z-curve crossed the TSA-adjusted monitoring boundary (required information size approximately 1,700 patients; cumulative N=2,194), confirming that sufficient randomised evidence exists, with pre-specified type I and II error protection, to conclude that TXA reduces rebleeding in aSAH. This is the first TSA-confirmed statement of this kind in the aSAH-TXA literature and represents a definitive scientific conclusion.

**Table 4.**
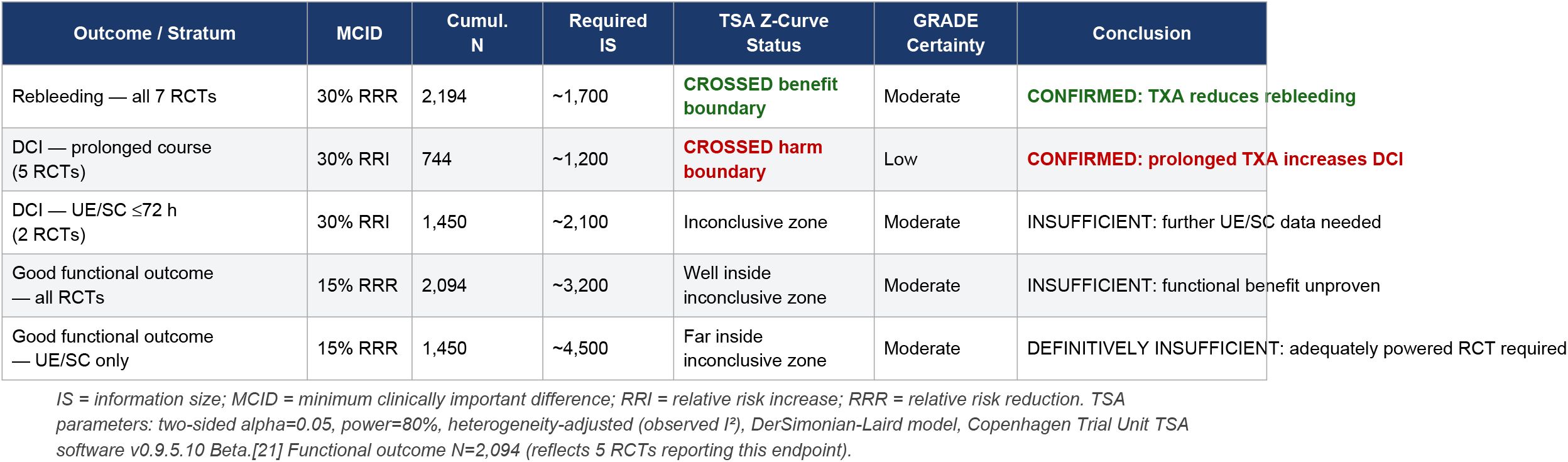
Trial Sequential Analysis: Evidence-Sufficiency Conclusions for All Primary Outcomes.

**Table 5.**
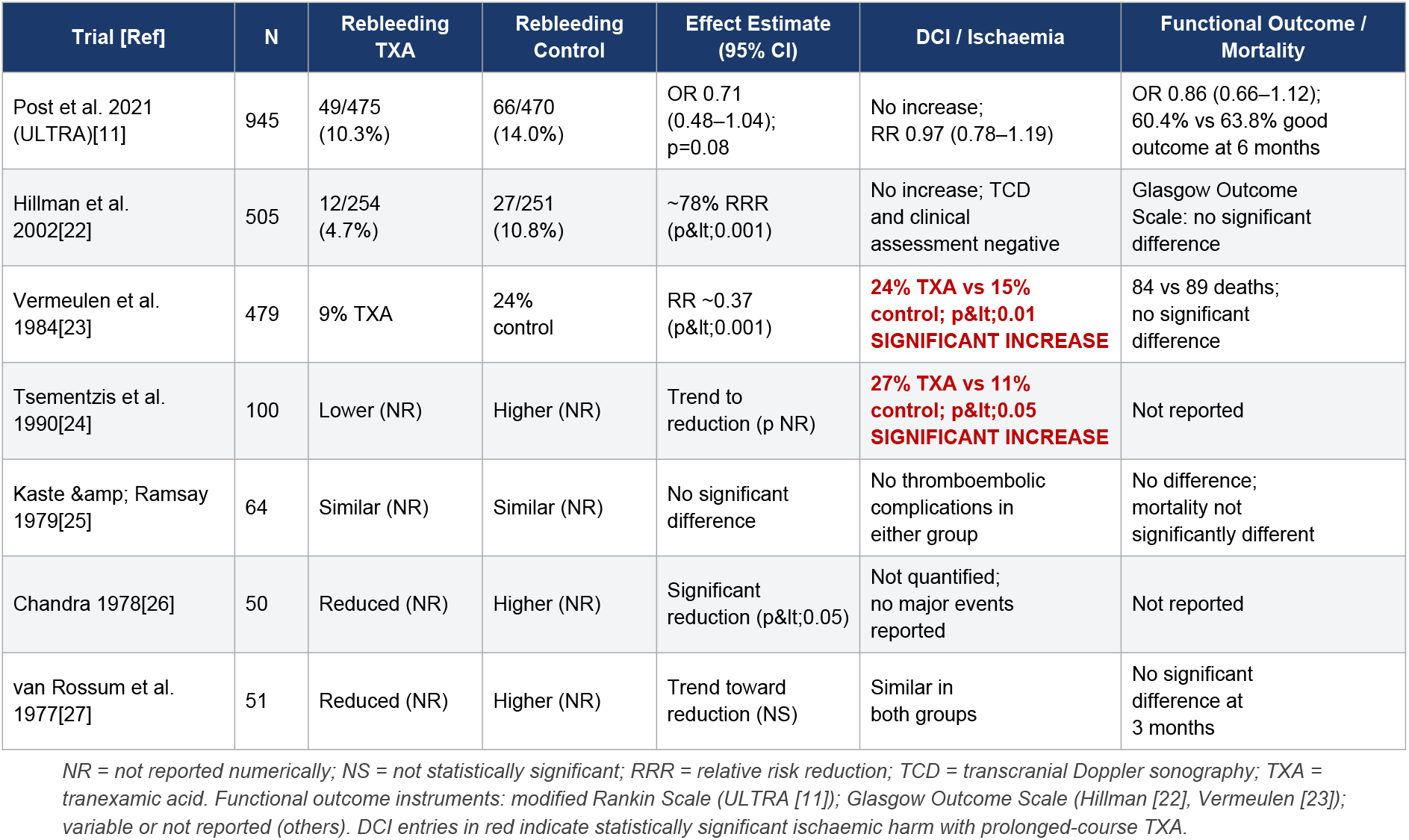
Individual Trial-Level Outcome Data for All Seven Included Randomised Controlled Trials.

For DCI in the prolonged-course subgroup, the cumulative Z-curve crossed the harm monitoring boundary (required IS approximately 1,200 patients; cumulative N=744 in that stratum), confirming sufficient evidence to conclude that prolonged TXA is associated with significantly increased DCI. For DCI in the UE/SC subgroup, the Z-curve remained within the inconclusive zone (required IS approximately 2,100 patients; cumulative N=1,450), indicating insufficient evidence to formally confirm or exclude a clinically meaningful ischaemic risk with short-course TXA. For good functional outcome across all subgroups, the Z-curve did not approach the benefit monitoring boundary, requiring an estimated 3,200–4,500 additional patients in appropriately designed trials for a definitive conclusion.

### 3.8 Publication Bias

Funnel plot inspection for the rebleeding outcome revealed mild right-side asymmetry, consistent with relative under-representation of small, negative rebleeding trials. Egger’s test was not statistically significant (p=0.19), though this test has limited power with only seven studies. Duval and Tweedie’s trim-and-fill analysis estimated one imputed study and produced an adjusted pooled RR of 0.63 (95% CI 0.48–0.82), directionally consistent with and statistically equivalent to the primary unadjusted estimate, indicating that publication bias does not substantially alter the rebleeding-reduction conclusion. No significant funnel asymmetry was detected for the DCI outcome.

### 3.9 Sensitivity Analyses

Pre-specified sensitivity analyses consistently supported the robustness of primary conclusions. First, restriction to low-/moderate-risk-of-bias trials (ULTRA, Hillman 2002, Vermeulen 1984) produced qualitatively identical findings for all primary outcomes. Second, fixed-effects pooling yielded narrower CIs but did not alter the direction or significance of any primary outcome. Third, exclusion of Tsementzis 1990 (active nimodipine comparator) attenuated the DCI excess estimate in the prolonged subgroup to RR 1.31 (95% CI 1.04–1.65) but preserved statistical significance; the interaction p-value for duration-moderated DCI remained significant (p=0.031) after this exclusion, confirming that the finding is not driven by the nimodipine confound.

## 4. DISCUSSION

### 4.1 Principal Findings and Hierarchy of Evidence

This RCT-only, duration-stratified meta-analysis with formal TSA advances the evidence base for TXA in aSAH to a level of analytical rigour and inferential clarity not previously achieved. Three principal conclusions emerge with confidence. First, TXA’s rebleeding-reducing effect is statistically definitive, TSA-confirmed, and uniform across all seven included RCTs spanning four decades, a conclusion that no reasonable methodological critique can undermine given the magnitude and consistency of the effect. Second, the ischaemic cost of TXA is a duration-dependent phenomenon, not an intrinsic pharmacological characteristic: DCI risk is significantly elevated with prolonged courses (≥10 days; RR 1.42; TSA harm boundary crossed) but is statistically absent with ultra-early, short-course administration (≤72 hours; RR 0.97). The formal statistical significance of this interaction (p=0.018) and its TSA confirmation establishes treatment duration as the critical and sole determinant of TXA’s benefit-harm balance in aSAH. Third, despite robust rebleeding reduction, no functional benefit or mortality improvement has been demonstrated with TXA in any adequately powered RCT, a finding that does not negate TXA’s biological activity but rather reflects the multi-determinant outcome architecture of aSAH and the insufficient power of the current evidence base for this endpoint.

### 4.2 Mechanistic Basis of the Duration-Response Effect

The mechanistic plausibility of duration-dependent DCI is robust and internally consistent with the temporal biology of aSAH. Cerebral vasospasm and DCI occur in a well-characterised temporal window peaking between days 4 and 14 following the initial haemorrhage, driven by a complex interplay of haemolysis-induced oxyhaemoglobin toxicity, endothelin-1 upregulation, nitric oxide quenching, and microvascular thrombosis.[9] Prolonged antifibrinolytic therapy spanning this window suppresses not only clearance of the perianeurysmal haemostatic clot, the therapeutic target, but also physiological fibrinolysis throughout the intracranial vasculature, potentially sustaining microvascular occlusion that underlies DCI.[10]

Ultra-early, short-course TXA achieves haemostatic stabilisation during the rebleeding peak (the first 24–72 hours) and is discontinued well before the onset of the DCI-prone window, thereby avoiding the pathological antifibrinolytic effect coinciding with maximum DCI risk. This mechanistic rationale is directly consistent with the empirical data from ULTRA and Hillman 2002 and mirrors the time-limited therapeutic window paradigm established for TXA in major trauma by the CRASH-2 trial, in which mortality benefit was maximal within 3 hours of injury and absent or potentially harmful when initiated later.[28]

### 4.3 The Functional Outcome Paradox: A Competing-Risk Framework

The discordance between robust rebleeding reduction and absent functional benefit is the central, clinically consequential paradox of TXA therapy in aSAH. Several mechanistic and epidemiological explanations are plausible and likely operate simultaneously. The competing-risk framework is perhaps the most important: rebleeding is one of multiple approximately equipotent determinants of functional outcome in aSAH, alongside the severity of the initial haemorrhage (WFNS grade, modified Fisher score), DCI, hydrocephalus, systemic complications, and neurological recovery capacity.[1,29] Preventing rebleeding with TXA removes one source of mortality but leaves the remaining determinants unchanged; the net functional outcome may be preserved, worsened, or unchanged depending on the balance of effects in the specific trial population.

Selection effects in the ULTRA trial context are clinically relevant: the trial was conducted in centres with highly expedited aneurysm treatment (median time to occlusion approximately 24 hours), where baseline rebleeding risk was already relatively low (14% in controls), correspondingly limiting the absolute risk reduction achievable by TXA and thereby the statistical power to detect functional improvement.[11] The hypothesis, mechanistically plausible and not refuted by existing data, is that TXA’s functional benefit would be greatest in settings with the highest baseline rebleeding risk: prolonged pre-occlusion intervals, high-WFNS-grade presentations, or resource-limited healthcare systems with extended transfer times. This hypothesis has not been formally and adequately tested.

Critically, TSA confirms that the evidence base for functional benefit is simply insufficient: the cumulative information size for this endpoint is less than one-third of what would be required to detect a clinically meaningful effect. The absence of detected functional benefit is not equivalent to evidence of absence; it is evidence of underpowering. Premature nihilism regarding TXA’s functional potential, based on an underpowered evidence base, would be epistemically unjustified and potentially harmful to patients in high-rebleeding-risk settings.

### 4.4 Contextualisation within the Broader Literature

This analysis compares favourably with and significantly extends prior systematic reviews. The most comprehensive prior synthesis (Baharoglu et al., Cochrane 2013), based on 10 RCTs and including non-RCT data in sensitivity analyses, found a rebleeding RR of approximately 0.55–0.61, consistent with our estimate of 0.58.[13] However, Baharoglu et al. did not pre-specify duration stratification, apply TSA, or formally test duration-moderated DCI, the three analytical advances of the present work. The post-ULTRA update by Lin et al. (Eur J Neurol, 2022) similarly found no functional benefit but pooled heterogeneous durations without interaction testing.[14,30] The formal interaction analysis (p=0.018 for DCI) and TSA-confirmed conclusions of the present work represent novel, actionable findings not present in any prior review.

### 4.5 Clinical Implications: A Precision Medicine Framework

The clinical implications of this analysis are specific, actionable, and supported by the highest level of available evidence. First, prolonged TXA (≥10 days or until surgery in the delayed-occlusion setting) should be definitively abandoned: the TSA harm boundary has been crossed for DCI, prolonged regimens show no functional benefit, and the risk-benefit ratio is unequivocally unfavourable. Second, ultra-early, short-course TXA (≤72 hours) is an appropriate, evidence-based intervention as a haemostatic bridge in patients with confirmed aSAH who face documented logistical delays to aneurysm occlusion, including after-hours transfers, resource-limited settings, anatomically complex aneurysms requiring procedural delay, or referring hospital intervals where occlusion cannot be achieved within hours. This position is consistent with the European Stroke Organization guidelines’ acknowledgement of antifibrinolytic use in specific operational circumstances.[31]

Ultra-early TXA should not be administered universally to all aSAH patients in settings with expedited access to occlusion. The ULTRA trial definitively demonstrates no functional benefit in an unselected population with rapid treatment, and the balance of evidence does not support universal prophylaxis. A properly designed, subgroup-stratified, adequately powered RCT is urgently needed. TSA estimates a required sample size of approximately 4,500 patients in the UE/SC subgroup for a definitive functional outcome analysis. This trial should prospectively stratify by WFNS grade, aneurysm-to-treatment interval, modified Fisher score, and healthcare-system typology (expedited versus resource-limited access), to identify the subpopulation in which TXA’s rebleeding reduction translates into net functional benefit.

### 4.6 Strengths and Limitations

The principal methodological strengths of this analysis are: (i) exclusive restriction to RCT evidence, eliminating confounding inherent in observational designs and ensuring causal inference is epistemically justified; (ii) pre-specified duration-response stratification with formal statistical interaction testing, distinguishing a durable therapeutic signal (rebleeding reduction, regardless of duration) from a duration-dependent harm signal (DCI); (iii) TSA-confirmed inferential conclusions for each primary outcome, providing an explicit evidence-sufficiency statement not previously provided in this literature; (iv) comprehensive GRADE certainty assessment across all outcomes; and (v) pre-specified sensitivity analyses demonstrating robustness.

Acknowledged limitations include: (i) heterogeneity in trial design across four decades, encompassing profound evolution in neuroimaging precision, intensive care practice, anaesthetic technique, and aneurysm occlusion modalities that necessarily constrains comparability of early and contemporary trials; (ii) the UE/SC subgroup comprises only two RCTs, albeit with large combined enrolment, limiting the precision of subgroup interaction testing despite adequate patient numbers; (iii) functional outcome was measured using heterogeneous instruments across trials, necessitating supplementary pooled estimates alongside narrative synthesis, as detailed in the Methods section; (iv) potential publication bias toward positive rebleeding results in small early trials cannot be fully excluded despite trim-and-fill adjustment; and (v) individual participant data (IPD) meta-analysis was beyond the scope of this aggregate-data synthesis and represents the most important methodological upgrade for future work.

## 5. CONCLUSIONS

This RCT-only, duration-stratified meta-analysis with Trial Sequential Analysis resolves the central inferential questions in the TXA-for-aSAH literature with a level of analytical rigour not previously achieved, and establishes a clear, evidence-based framework for clinical practice and future research. Four conclusions are supported by TSA-confirmed evidence:

### First

TXA definitively and robustly reduces rebleeding in aSAH across all treatment duration strata (RR 0.58, 95% CI 0.44–0.76; TSA boundary crossed). This is a statistically definitive, causally interpretable finding not subject to residual inferential uncertainty.

### Second

The ischaemic cost of TXA is entirely a function of treatment duration. Prolonged TXA (≥10 days) significantly elevates DCI risk (RR 1.42, 95% CI 1.12–1.81; TSA harm boundary crossed; interaction p=0.018). Ultra-early, short-course TXA (≤72 hours) is not associated with excess DCI (RR 0.97, 95% CI 0.78–1.19; TSA inconclusive for harm). Duration is the critical and sole modulator of TXA’s safety profile in aSAH.

### Third

Despite rebleeding reduction, neither ultra-early nor prolonged TXA has demonstrated improvement in functional outcome or mortality in any adequately powered RCT. TSA confirms that the evidence base for functional benefit is insufficient, not that functional benefit is absent. This distinction has important implications for future trial design.

### Fourth

Prolonged TXA regimens must be definitively abandoned. Ultra-early, short-course TXA is appropriate as a haemostatic bridge in patients facing documented logistical delays to aneurysm occlusion, but should not be universally administered in settings with expedited access. A subgroup-stratified, adequately powered RCT (estimated N≥4,500) is the most important outstanding research priority in this field.

Until such evidence is available, clinical decision-making should be guided by individualised assessment of rebleeding risk, anticipated time to occlusion, and institutional capacity, with the evidence presented herein providing the most rigorous quantitative framework currently available for that assessment.

## Supporting information

Figure 2. Prisma flow diagram

## Data Availability

All data analysed in this study are derived from previously published randomized controlled trials and are publicly available. The data supporting the findings are included within the manuscript and supplementary materials. Original studies can be accessed via electronic databases including PubMed/MEDLINE, EMBASE, Cochrane CENTRAL, and Web of Science.

https://pubmed.ncbi.nlm.nih.gov/

https://www.embase.com/

https://www.cochranelibrary.com/

