## Supplementary material for "Ultra-Early, Short-Course Tranexamic Acid in Aneurysmal Subarachnoid Haemorrhage: An RCT-Only Meta-Analysis of Rebleeding Prevention Versus Ischaemic Harm, with Duration-Response and Trial Sequential Analysis": Figure 2. Prisma flow diagram

### Figure 2. PRISMA 2020 Flow Diagram

*Systematic Review and Meta-Analysis: Tranexamic Acid in Aneurysmal Subarachnoid Haemorrhage*

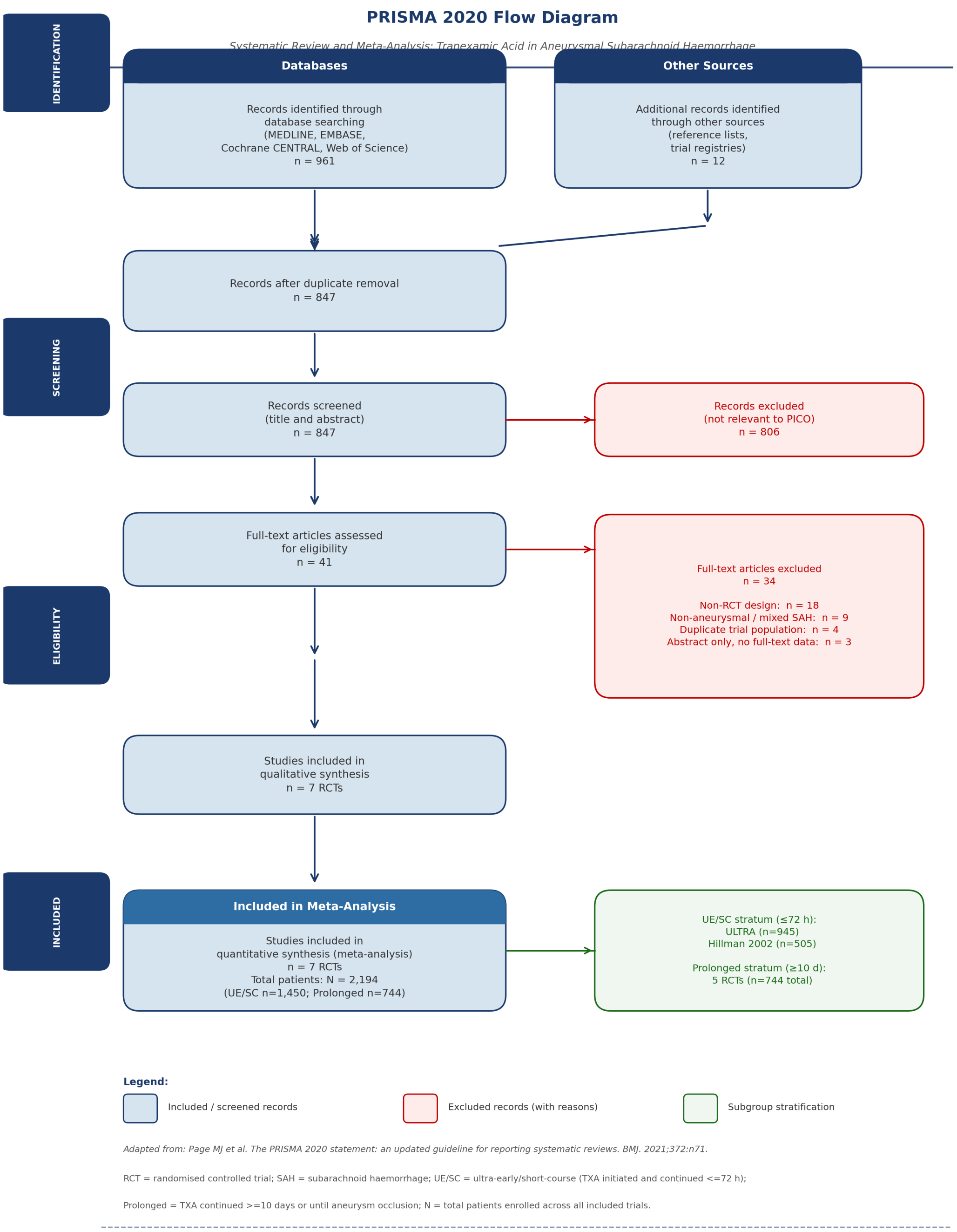


*Figure 2 legend. PRISMA 2020 flow diagram illustrating the complete systematic literature search and study selection process. A total of 961 records were identified through electronic database searching (MEDLINE, EMBASE, Cochrane CENTRAL, Web of Science) and 12*

*additional records through reference list and trial registry searches (ClinicalTrials.gov, WHO ICTRP). After removal of 126 duplicates, 847 unique records were screened by title and abstract. Eight hundred and six records were excluded as not relevant to the PICO question. Forty-one full-text articles were assessed for eligibility against pre-specified inclusion criteria; 34 were excluded with reasons as detailed (18 non-RCT design, 9 non-aneurysmal or mixed SAH, 4 duplicate trial populations, 3 abstract only without full-text data). Seven RCTs meeting all inclusion criteria were included in the qualitative and quantitative synthesis (total N = 2,194 patients). Subgroup stratification: ultra-early/short-course (UE/SC) stratum, TXA initiated and continued ≤72 hours (ULTRA n = 945; Hillman 2002 n = 505; stratum total n = 1,450); prolonged-course stratum, TXA continued ≥10 days or until aneurysm occlusion (five RCTs; stratum total n = 744). Adapted from: Page MJ et al. The PRISMA 2020 statement: an updated guideline for reporting systematic reviews. BMJ. 2021;372:n71. UE/SC = ultra-early/short-course; RCT = randomised controlled trial; SAH = subarachnoid haemorrhage; PICO = Population, Intervention, Comparator, Outcome.*
